# Does Vitamin D Supplementation Modulate Metabolic Risk Factors of Cardiovascular Disease? A Systematic Review and Meta-Analysis of Clinical Trials

**DOI:** 10.64898/2026.02.13.26346232

**Authors:** Suhad Abumweis, Sarah Alqadi, Ala’a Al Tarteer, Waed Alrefai, Foad Alzoughool, Stephanie Jew, Taima Qudah

**Affiliations:** Department of Clinical Nutrition and Dietetics, Faculty of Applied Medical Sciences, The Hashemite University, Zarqa,Jordan; Independent Researcher, Jordan; Department of Health Science and Biostatistics, Swinburne University of Technology, Hawthorn, VIC 3122, Australia; Department of Medical Laboratory Sciences, Faculty of Applied Medical Sciences, The Hashemite University, P.O. Box 330127, Zarqa 13133, Jordan; Faculty of Health Sciences, Fujairah Women’s College, Higher Colleges of Technology, UAE; Independent Researcher, Canada; College of Pharmacy and AAU Health and Biomedical Research Center, AL Ain University, Abu Dhabi, P.O.Box 112612, United Arab Emirates

**Author notes:** Stephanie Jew is currently a federal public servant, however, the opinion presented in this article are not those of the Government of Canada. **Corresponding Author:** Suhad Abumweis, PhD Department of Clinical Nutrition and Dietetics, Faculty of Applied Medical Sciences, The Hashemite University, Jordan.

**Keywords:** Vitamin D, cardiovascular disease, cardiometabolic risk factors, systolic blood pressure, low-density lipoprotein cholesterol

## Abstract

**Background:** Vitamin D supplementation has been investigated for potential associations with cardiometabolic risk factors related to cardiovascular disease (CVD); however, findings from randomized controlled trials (RCTs) remain inconsistent. This meta-analysis aimed to assess the effects of vitamin D supplementation on cardiometabolic risk factors—including lipid profile, blood pressure, and glycaemic parameters—and to explore whether age and baseline serum vitamin D concentrations modify these associations.

**Research Design and Methods:** We conducted a systematic review and meta-analysis of RCTs comparing oral vitamin D supplementation with placebo in adults. PubMed, the Cochrane Library, and ClinicalTrials.gov. Risk of bias was evaluated using the Cochrane tool, and pooled effect sizes with 95% confidence intervals (CIs) were calculated using random-effects models.

**Results:** 14,051 abstracts were retrieved, of which 45 were used for data analysis. Vitamin D supplementation reduced low-density lipoprotein cholesterol (LDL-C) by 0.136 mmol/L (95%CI: -0.215, -0.56), systolic blood pressure by 2.79 mm Hg (95% CI: -4.648, -0.938), fasting blood glucose by -0.11 (95%CI:-0.185, -0.036), and hemoglobin A1c by 0.164% (95%CI: -0.322, -0.006) compared with placebo. Subgroup analyses revealed reductions in SBP and LDL cholesterol among participants aged ≥55 years and reductions in fasting blood glucose in participants with age < 55 years. While favourable effects on fasting blood glucose and hemoglobin A1c were observed with a baseline blood level of vitamin D of concentrations (<50 nmol/L).

**Conclusions:** Vitamin D supplementation may be associated with modest modifications in selected cardiometabolic risk factors; including systolic blood pressure, LDL-cholesterol, fasting blood glucose, and hemoglobin A1c. Age and baseline vitamin D status appear to modulate these effects. The clinical relevance of these modest effects remains uncertain. Well-designed RCTs with standardized protocols are required to clarify potential effect modification by age and baseline vitamin D status.

**Trial Registration:** PROSPERO (CRD42020165293)

**Funding:** This research received funding from the Hashemite University, P.O. Box 330127, Zarqa 13133, Jordan

## Introduction

Cardiovascular disease remains the leading cause of death in developed and developing countries. ^1–3^ Consequently, there is growing interest in dietary supplements that may favorably influence both traditional and emerging risk factors for cardiovascular disease (CVD).

Several prospective human studies have demonstrated an association between vitamin D status and cardiovascular disease mortality risk. For instance, Ginned *et al*. ^4^ followed 3,408 participants for about 7.3 years and found that subjects with vitamin D levels less than 25.0 nmol/L were at higher risk for cardiovascular disease mortality (hazard ratio=2.36, 95% CI=1.17-4.75) than those with levels of 100 nmol/L or higher. Similarly, Doping *et al*.^5^ reported higher cardiovascular mortality among participants in the lowest two quartiles of vitamin D status (median concentrations of 7.6 and 13.3 ng/mL), with hazard ratios of 2.22 and 1.82, respectively, compared with participants in the highest quartile (median 28.4 ng/mL).

Vitamin D receptors are expressed in almost all human cells, supporting a potential role for vitamin D in multiple physiological processes relevant to cardiovascular health. Evidence suggests that vitamin D may influence several risk factors associated with cardiovascular disease including hypertension, hyperlipidemia, diabetes mellitus, inflammation, and endothelial dysfunction. Proposed mechanisms include modulation of renin-angiotensin-aldosterone systems, decrease in parathyroid hormones, enhancement in insulin secretion and insulin sensitivity, downregulation of inflammatory cytokines, and increase arterial intimae thickness.^6^ On the basis of these proposed biological mechanisms, numerous randomized clinical trials have been undertaken to evaluate the effect of vitamin D supplementation on metabolic risk factors associated with cardiovascular disease, however, mixed results have been reported. Therefore, pooling the results from these trials will help to evaluate the relationship between vitamin D supplements and metabolic risk factors associated with cardiovascular disease. Moreover, utilizing sub-group techniques to study multiple potential effect modifiers —such as age and baseline vitamin D levels— may help clarify the relationship between vitamin D supplements and metabolic risk factors for CVD, and explain observed heterogeneity, as suggested by previous meta-analyses ^7,8^

Accordingly, the objectives of this systematic review and meta-analysis are to (1) quantify the effect of vitamin D supplements on CVD metabolic risk factors including blood lipids [total cholesterol, low-density lipoprotein cholesterol (LDL-cholesterol), high-density lipoprotein cholesterol (HDL-cholesterol), triglycerides, blood pressure [systolic blood pressure (SBP), diastolic blood pressure (DBP), inflammation (C-reactive protein, interleukin-6), and parameters of glucose metabolism (fasting blood glucose, hemoglobin A1c%), body mass index (BMI), and (2) to examine whether age and baseline serum vitamin D concentrations modify the effects of vitamin D supplementation on these outcomes.

## Methods

### Registration and Reporting

The meta-analysis has been registered at PROSPERO (CRD42020165293). And it is reported accordance to the PRISMA (Preferred Reporting Items for Systematic Reviews and Meta-Analyses) checklist.

### Literature Search

Clinical trials were identified by searching the following databases PubMed, the Cochrane Library, and ClinicalTrial.gov from the commencement until March 2020. An updated search was carried out in PubMed from March 2020 to July 2024. The updated search resulted in inclusion of the 14 clinical trials in the final analysis.

For non-English language literature, studies were included if an English abstract provided sufficient data for extraction; otherwise, the study was excluded. A comprehensive search strategy was employed using the following terms and their combinations: *vitamin D*, *25-hydroxyvitamin D3*, *calcidiol*, *1,25-dihydroxyvitamin D*, *cholecalciferol (D3)*, and *ergocalciferol (D2)*.

### Eligibility Criteria and Study Selection

Trials were include in the analysis if they met the following criteria: 1) randomised control trials of parallel or crossover design, 2) participants were adults 3) they provided the intervention as vitamin D compared to a placebo, and 4) reported at least one outcome of interest, including lipid profile, blood pressure, inflammatory cytokines, and parameters of glucose metabolism. One reviewer conducted the initial screening of titles and abstracts, while a second reviewer independently verified the included and excluded studies.

### Risk of Bias Assessment

Randomised controlled studies were assessed for methodological quality with the Cochrane risk of bias tool^9^. Evaluating sequence generation, allocation concealment, blinding of participants and personnel, blinding of outcome assessors, incomplete outcome data, selective reporting, and other potential sources of bias. For each category, the risk of bias was then categorized as “low,” “high,” or “unclear” ^9^.

### Data Extraction

For eligible studies, data were extracted on mean values and standard deviations of outcomes, as well as study characteristics including trial design, intervention type, vitamin D dose (IU/day), duration of supplementation, and participant characteristics. Two reviewers independently extracted data, and a third reviewer cross-checked the extracted data. Disagreements were resolved through discussion.

The effect size for continuous outcomes was calculated as the difference in means and its standard error was calculated for every study using a specialized program (Comprehensive Meta-Analysis V2 (Biostat, Englewood, NJ, USA)) for calculating pooled effect size and 95% confidence interval (95%CI) and performing subgroup analysis by age group, and blood baseline vitamin D concentrations.

### Statistical Analysis and Management of Heterogeneity

The effect size for continuous outcomes was calculated as the mean difference with corresponding standard errors. Meta-analyses were conducted using Comprehensive Meta-Analysis software (version 2; Biostat, Englewood, NJ, USA).

Given the anticipated clinical and methodological heterogeneity among studies—including wide variation in vitamin D dose, supplementation duration, baseline vitamin D status, and participant health status—a random-effects model was applied initially for all pooled analyses. This approach accounts for both within-study and between-study variability and provides more conservative effect estimates when heterogeneity is present.

The heterogeneity or variation of the results across the trials was evaluated using the I^2^ statistic, which assesses the percentage of between-study variance due to study heterogeneity versus sampling error, ranging from 0.0% (no heterogeneity) to 100% (high heterogeneity). A guide to I^2^ interpretation is as follows: 0% to 40% might not be important; 30% to 60% may represent moderate heterogeneity; 50% to 90% may represent substantial heterogeneity; and 75% to 100%: considerable heterogeneity.

In addition, sensitivity analysis was carried out on all outcomes to explore the influence of each individual study on the estimated pooled effects and thus to check the robustness of findings in the presence of high heterogeneity.

The presence of publication bias was examined using a funnel plot. Subgroup analysis was used to explore the effects of potential modifiers on the outcomes of interest and data were presented as forest plots.

## Results

A total of 14,051 records were identified through database searching. After removal of duplicates and screening, 45 randomized controlled trials met the inclusion criteria and were used for data analysis (Figure 1). Studies were excluded for reasons including: methodological papers, did not measure any of the outcomes of interest, presented the results of the screening procedure, reported outcome after treatment cessation, intravenous infusion, a re-analysis of a previous report or results reported previously, assessed treatment efficacy and not effectiveness, no placebo or not appropriate control, could not find the full article, i.e., only abstract available or no access, participants were children or adolescents and in vitro study.

**Figure 1.**
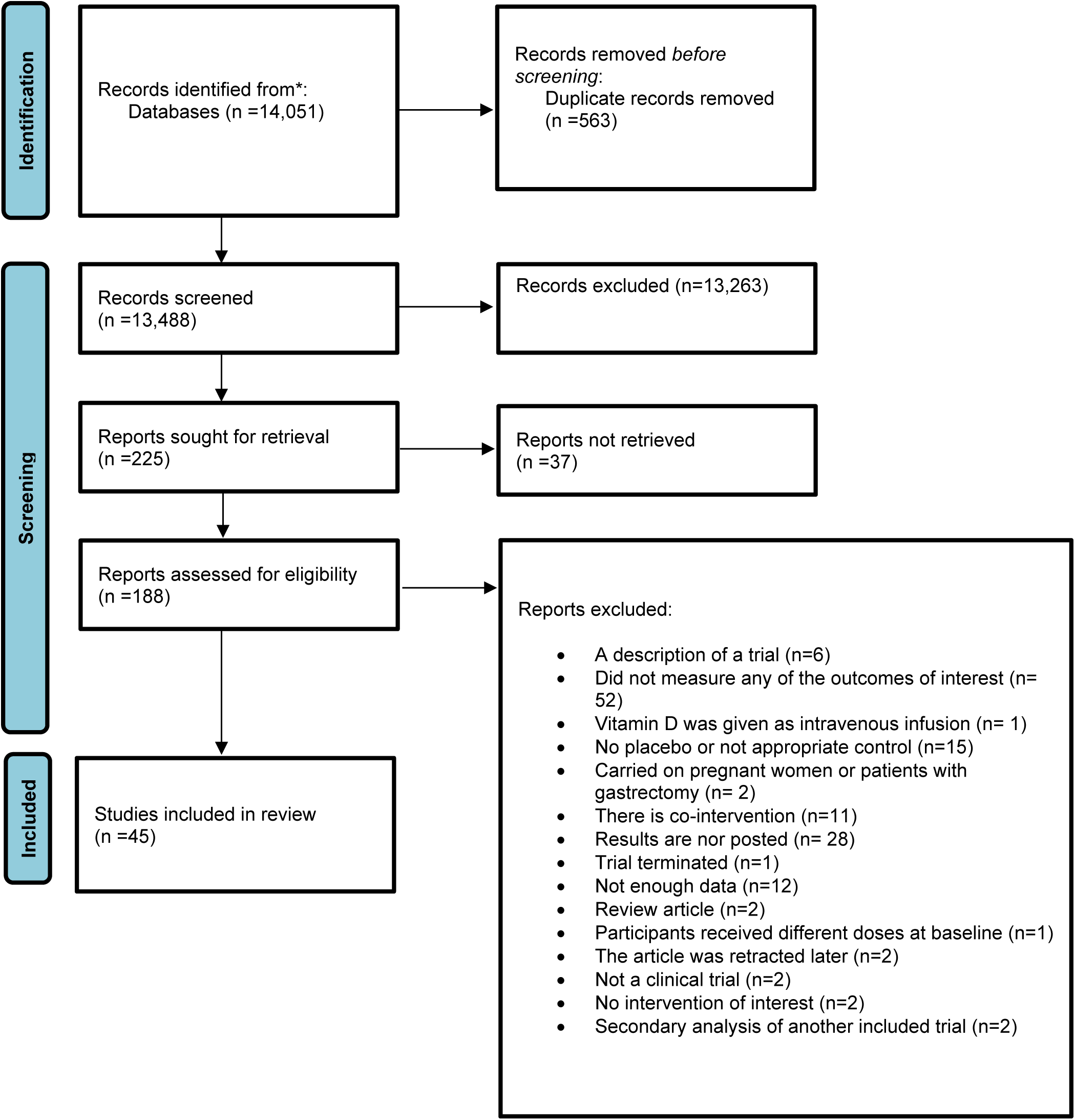
Flowchart of literature search for vitamin D supplementation and cardiovascular disease risk factors

Table 1 summarizes the characteristics of the included studies. All the studies were randomised, double-blinded with parallel design. study duration ranged from 1 to 36 months. Vitamin D dose ranged from 20 to 100,000 IU administered as daily, weekly, or bolus regimens. Most studies included both male and female participants. The majority of the studies supplied vitamin D in the form of cholecalciferol. Participants ranged in age from 18 to 88 years and included healthy individuals as well as populations with cardiovascular disease, diabetes, hypertension, overweight, obesity, coronary artery disease, polycystic ovary syndrome, chronic kidney disease, chronic heart failure, gestational diabetes mellitus, hypercholesterolemia, left ventricular hypertrophy, secondary hyperparathyroidism, ischemic heart disease, chronic fatigue, heart failure, metabolic syndrome, myocardial infarction, high waist circumference. Blood vitamin D concentration was assessed by using different laboratory methods such as enzyme-linked immunosorbent assay and liquid chromatography–tandem mass spectrometry. Studies evaluated mostly the efficacy of vitamin D3 supplement as a modulator of risk factors associated with cardiovascular disease.

**Table 1:**
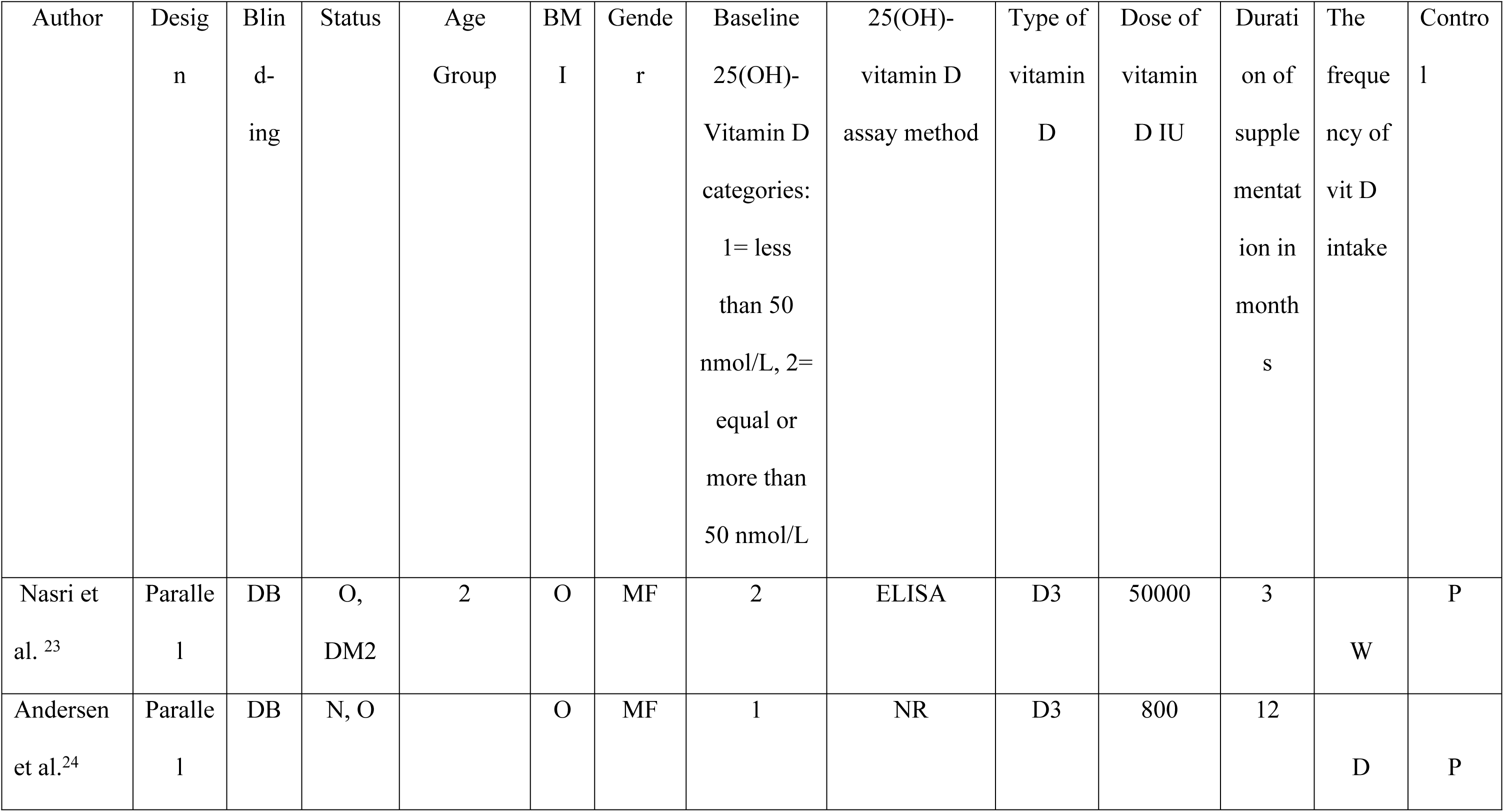
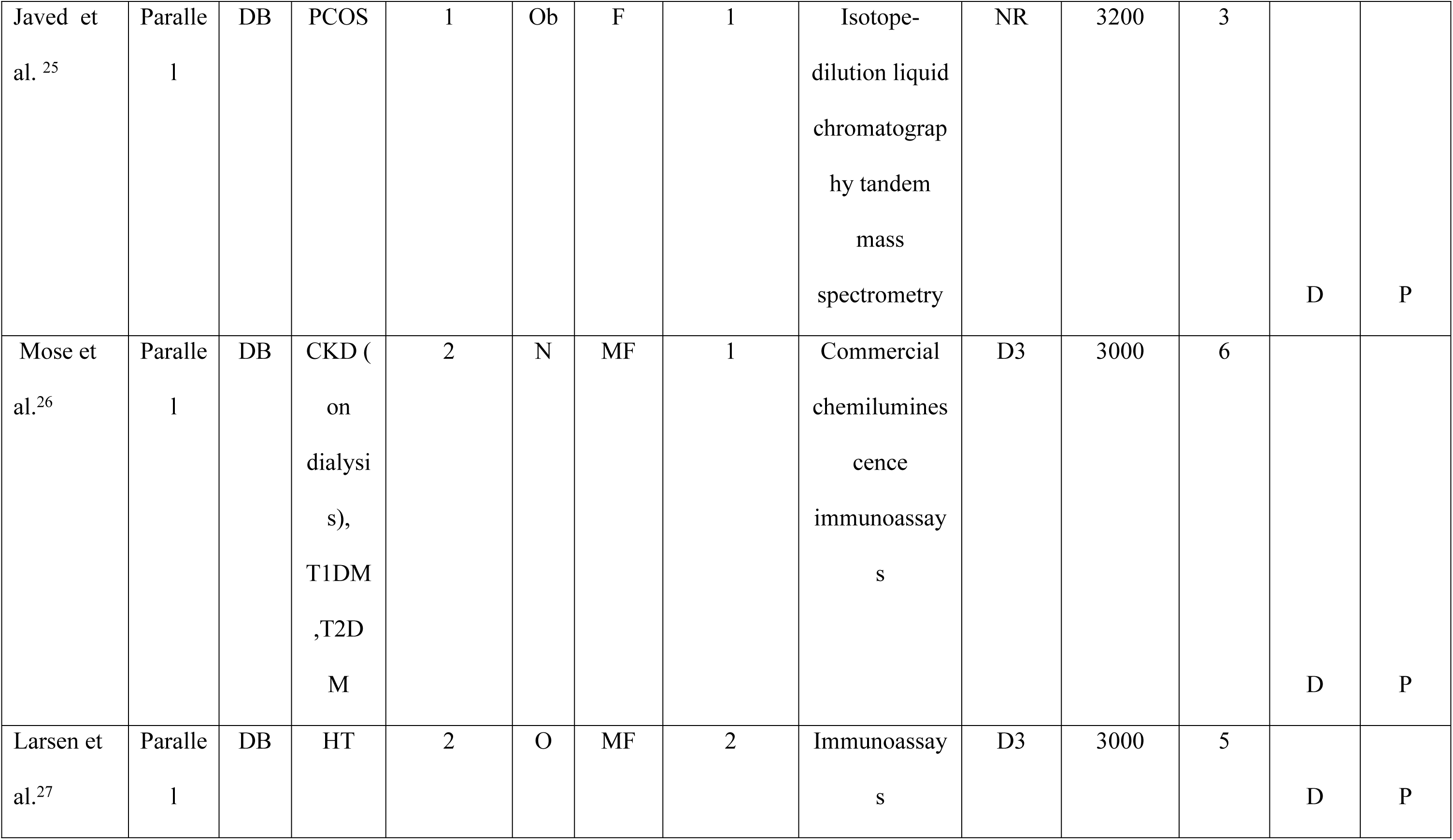
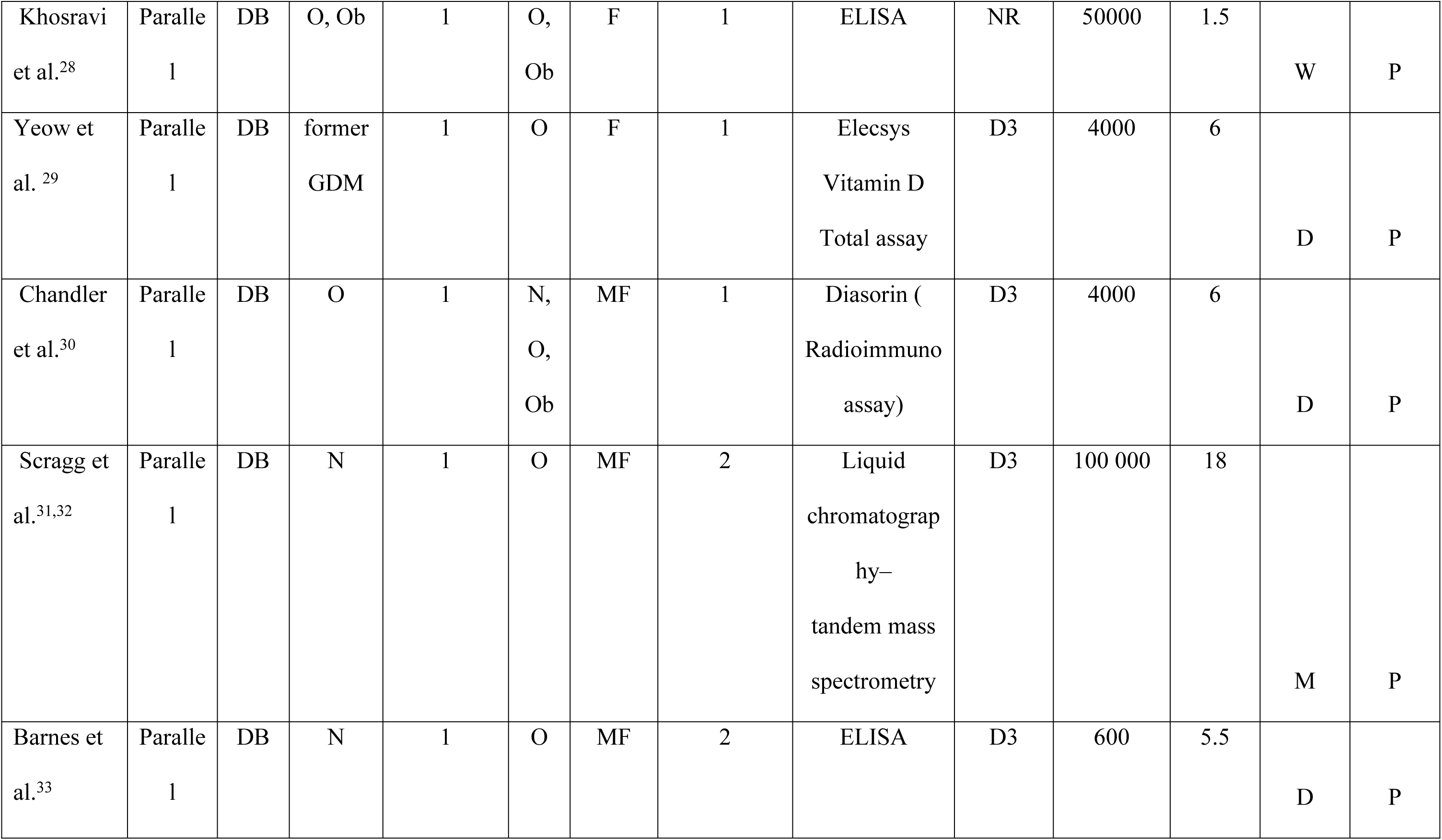
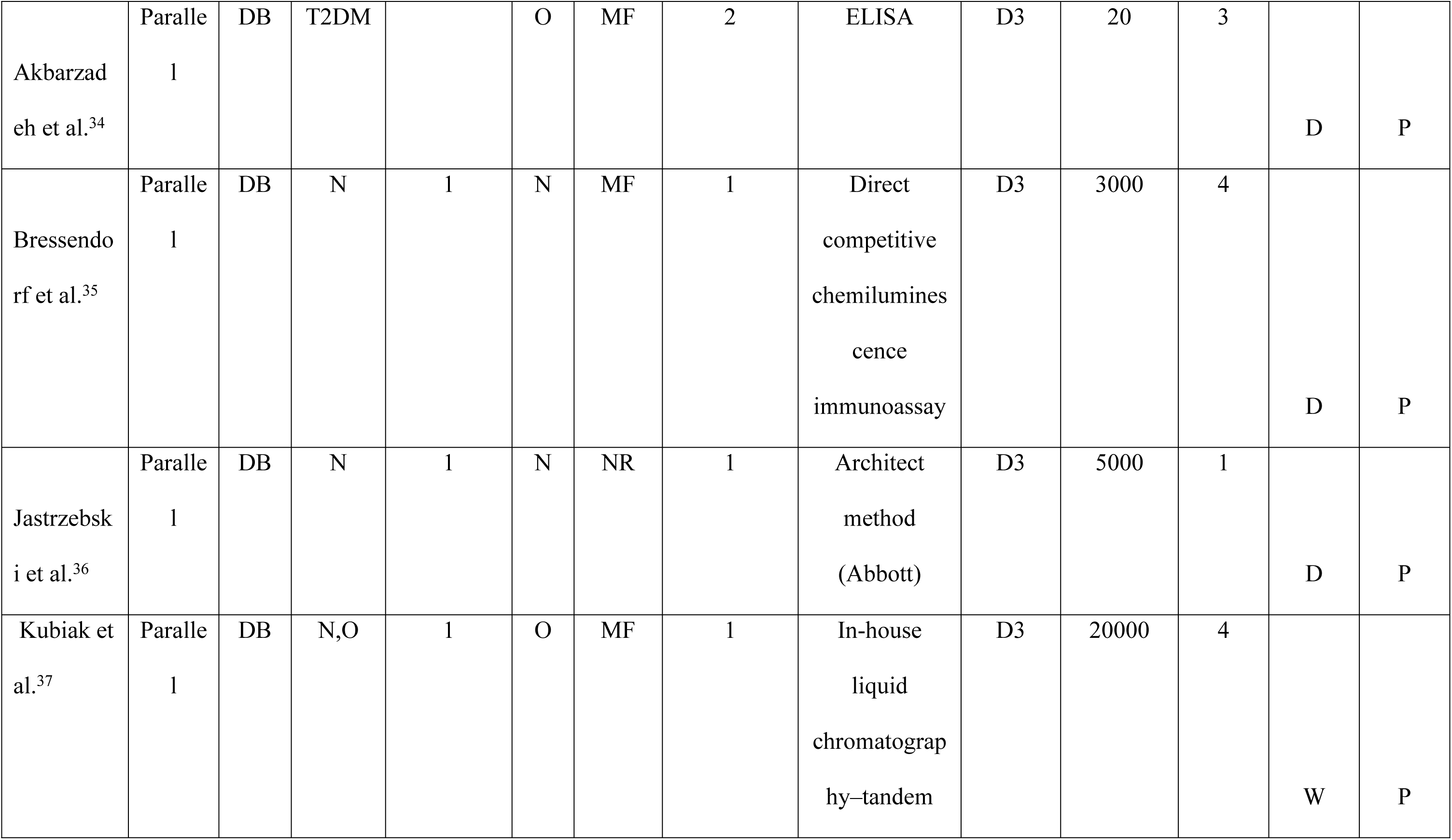
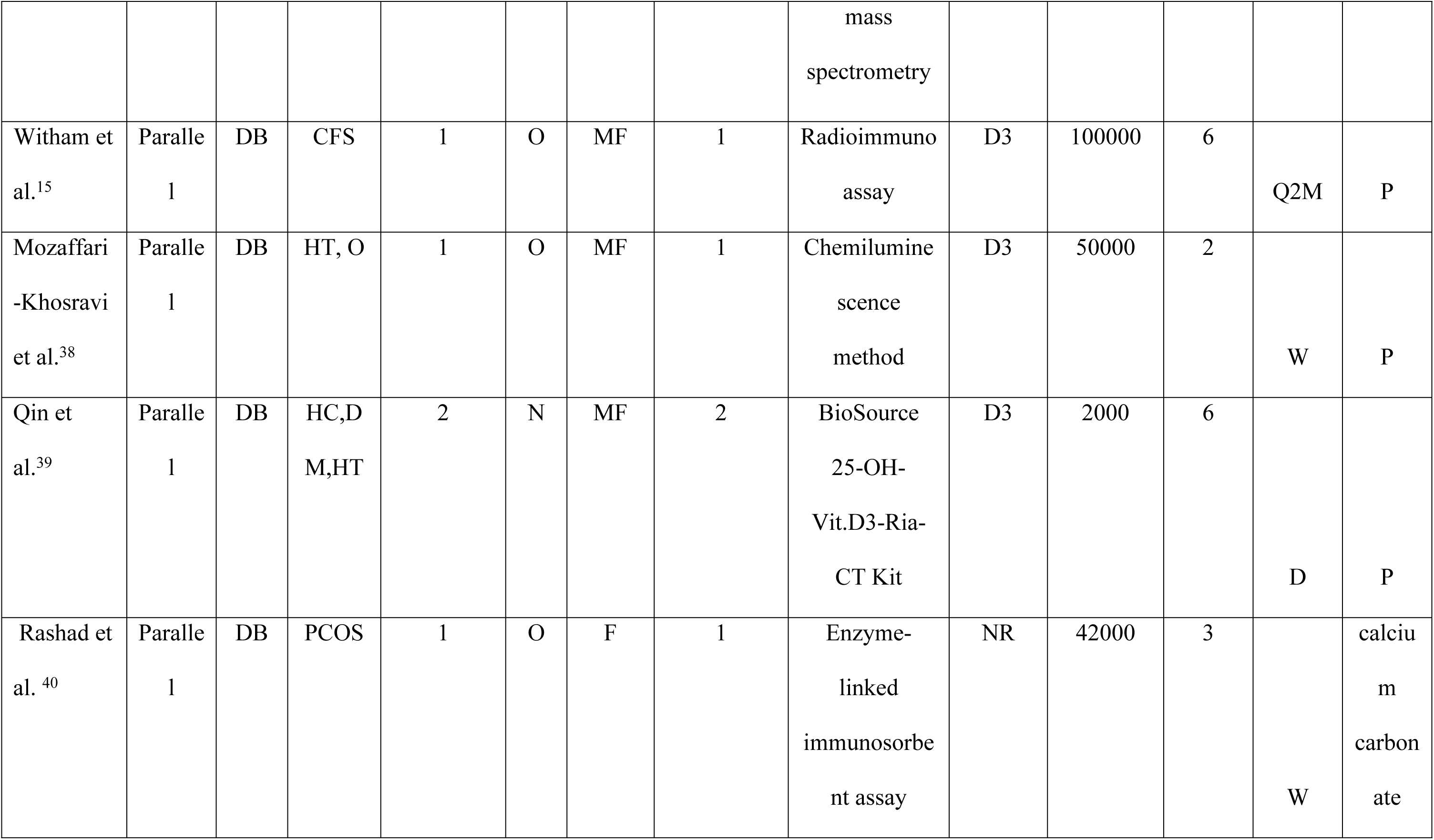
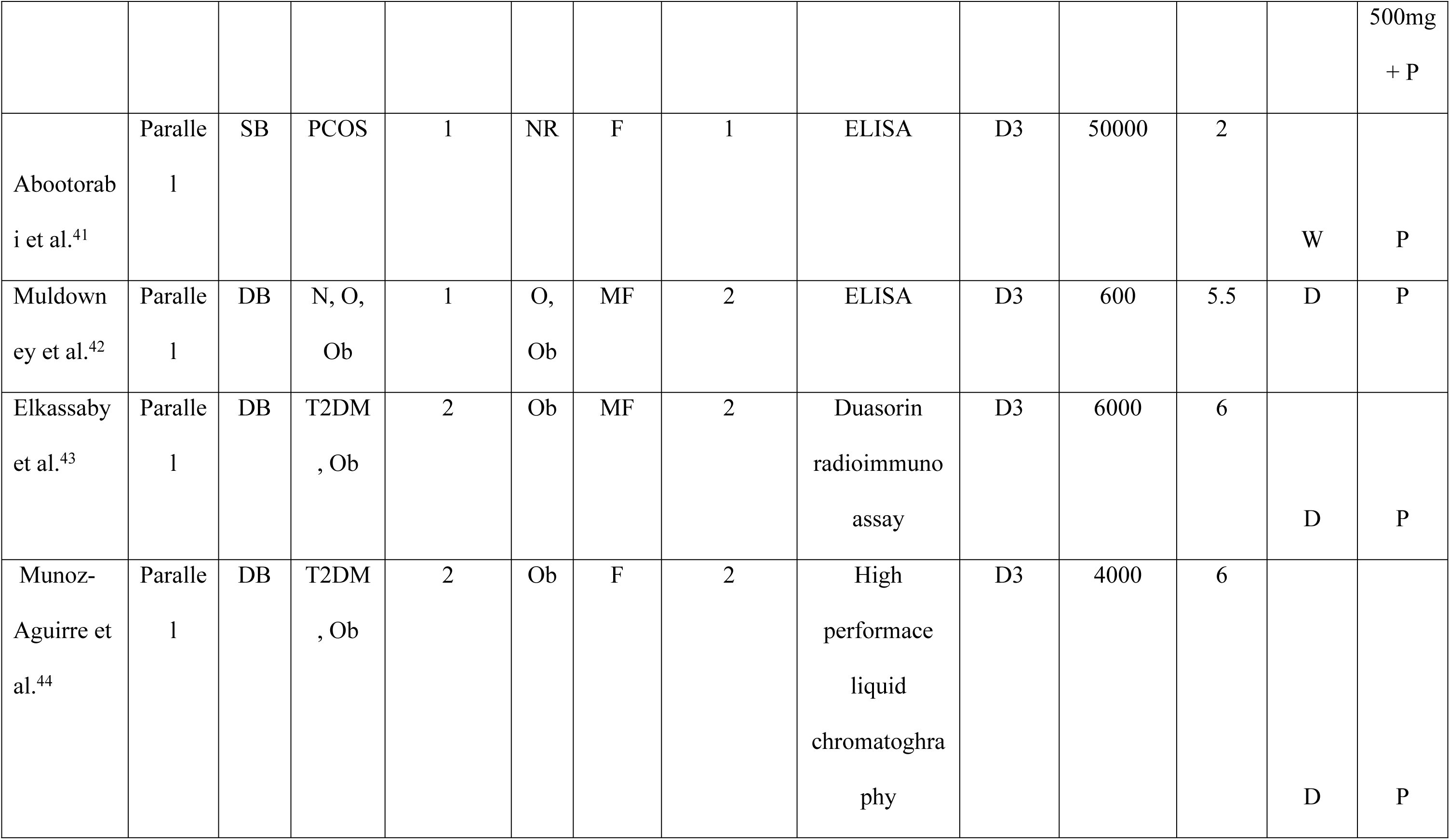
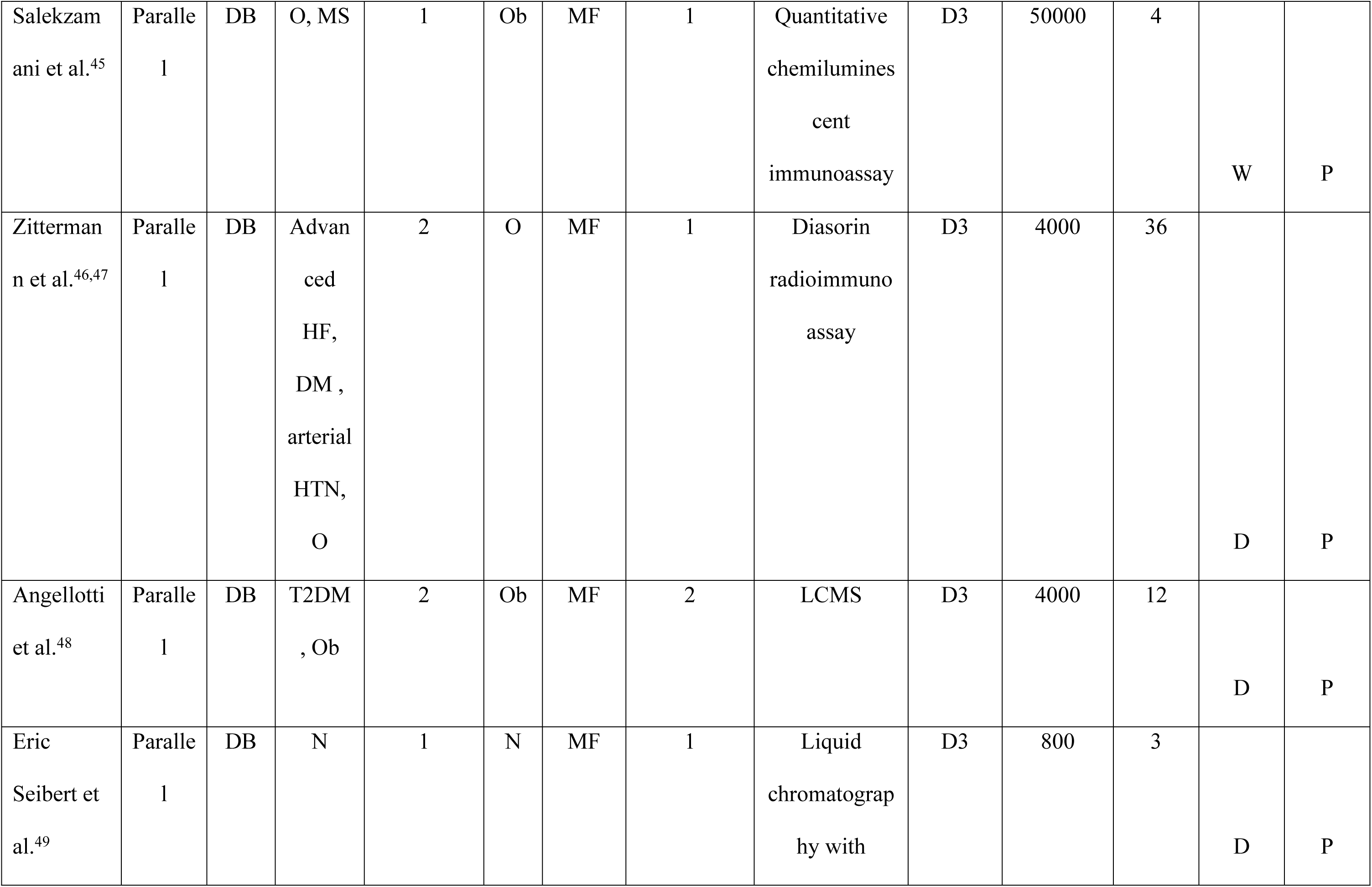
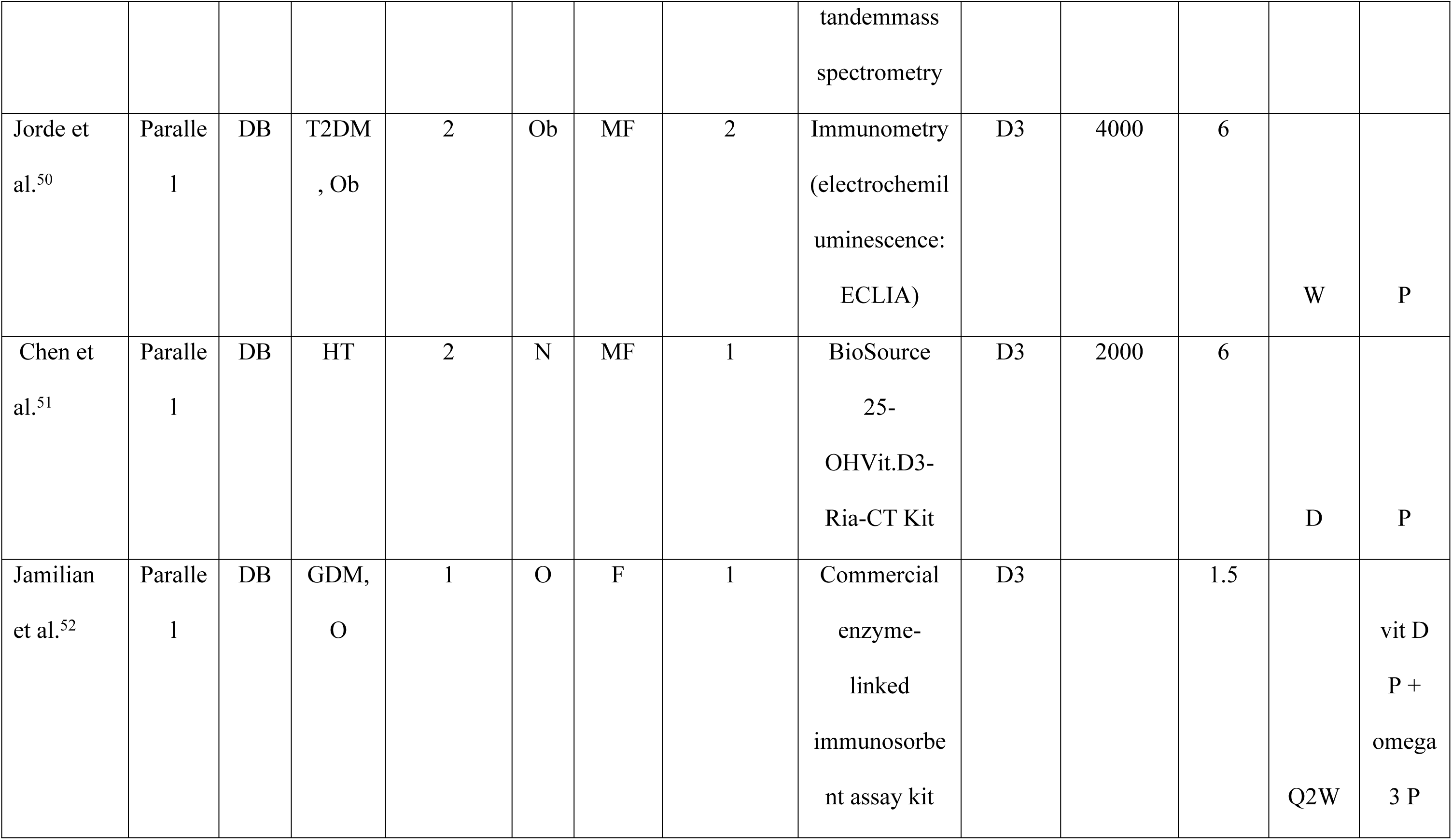
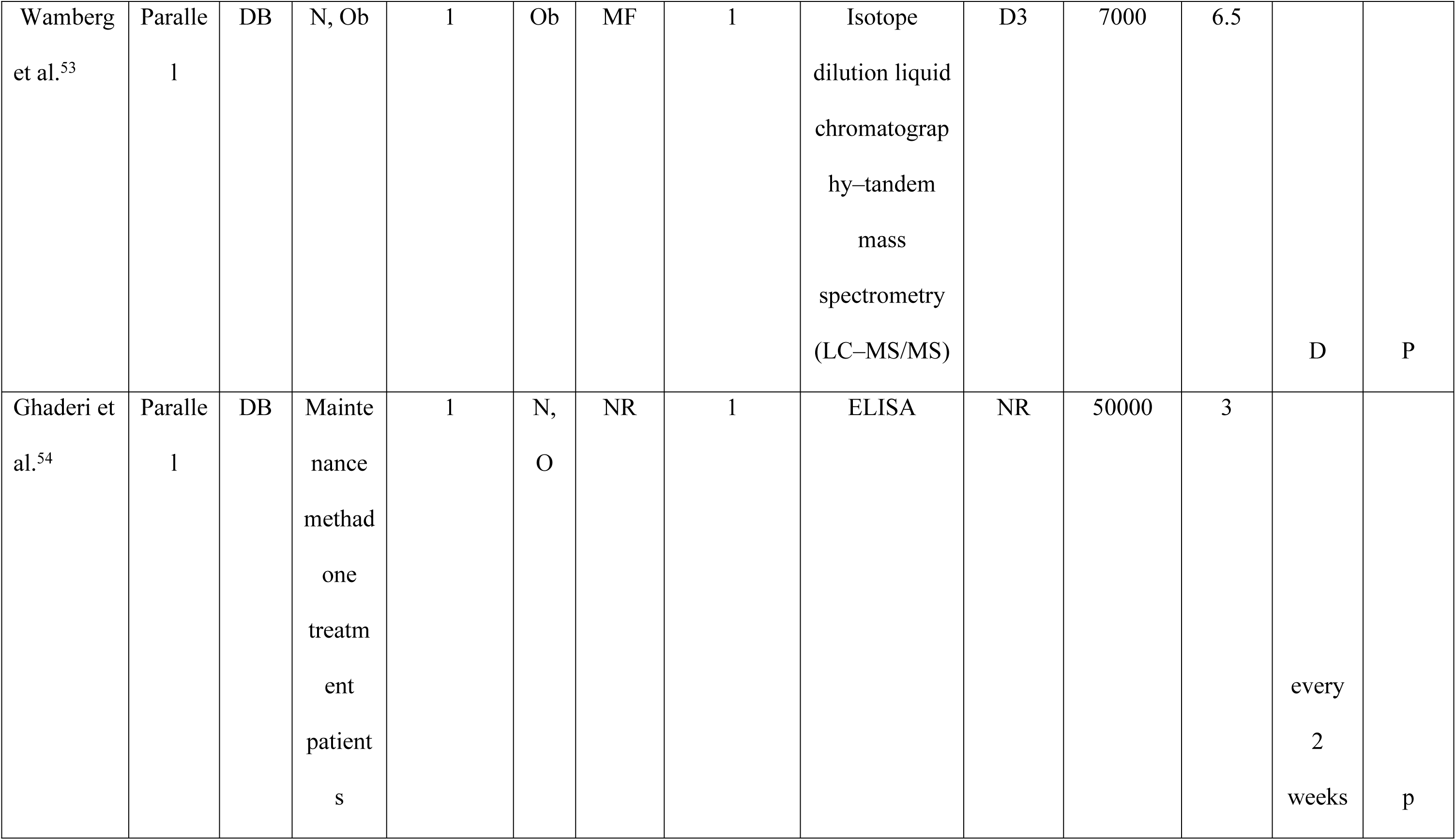
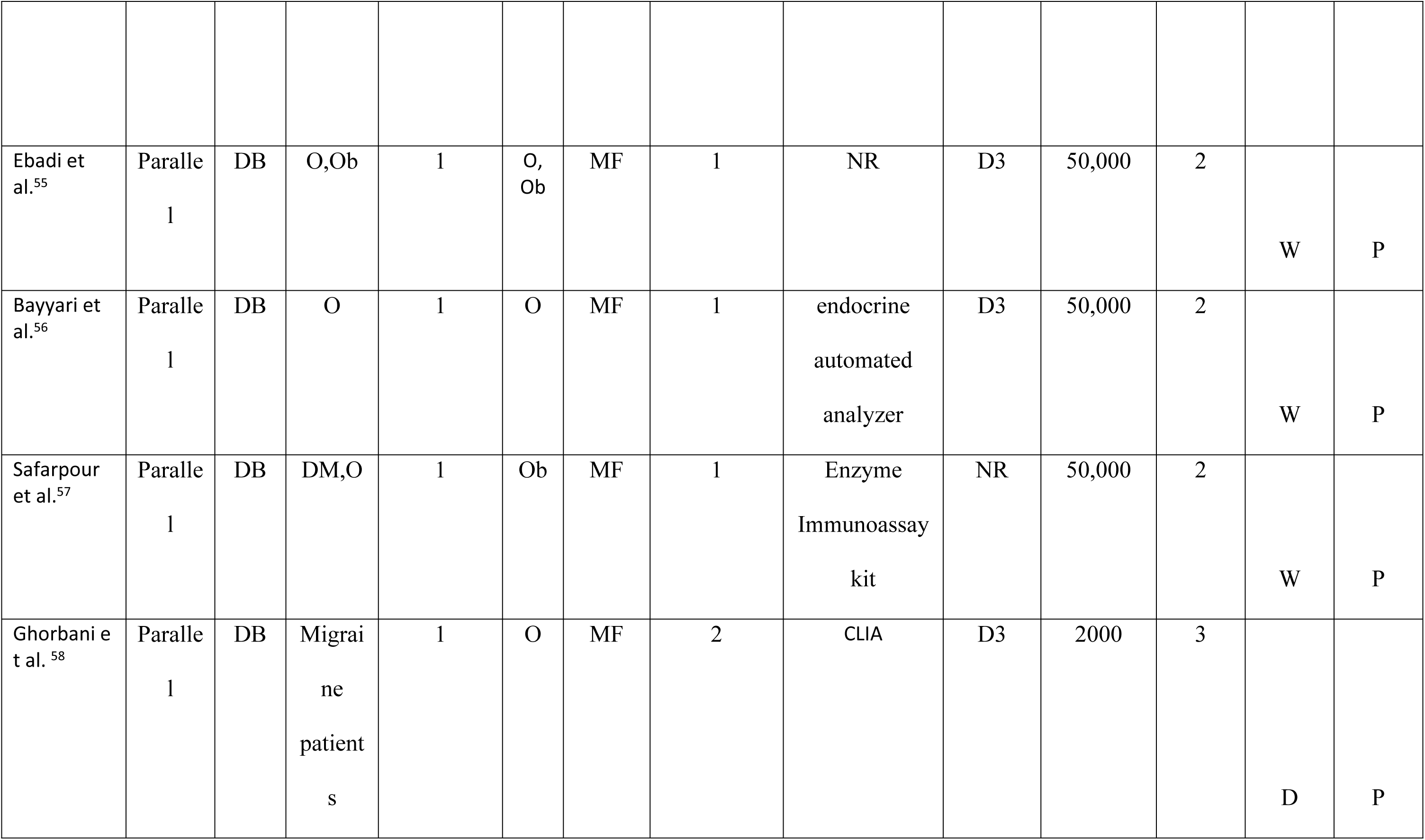
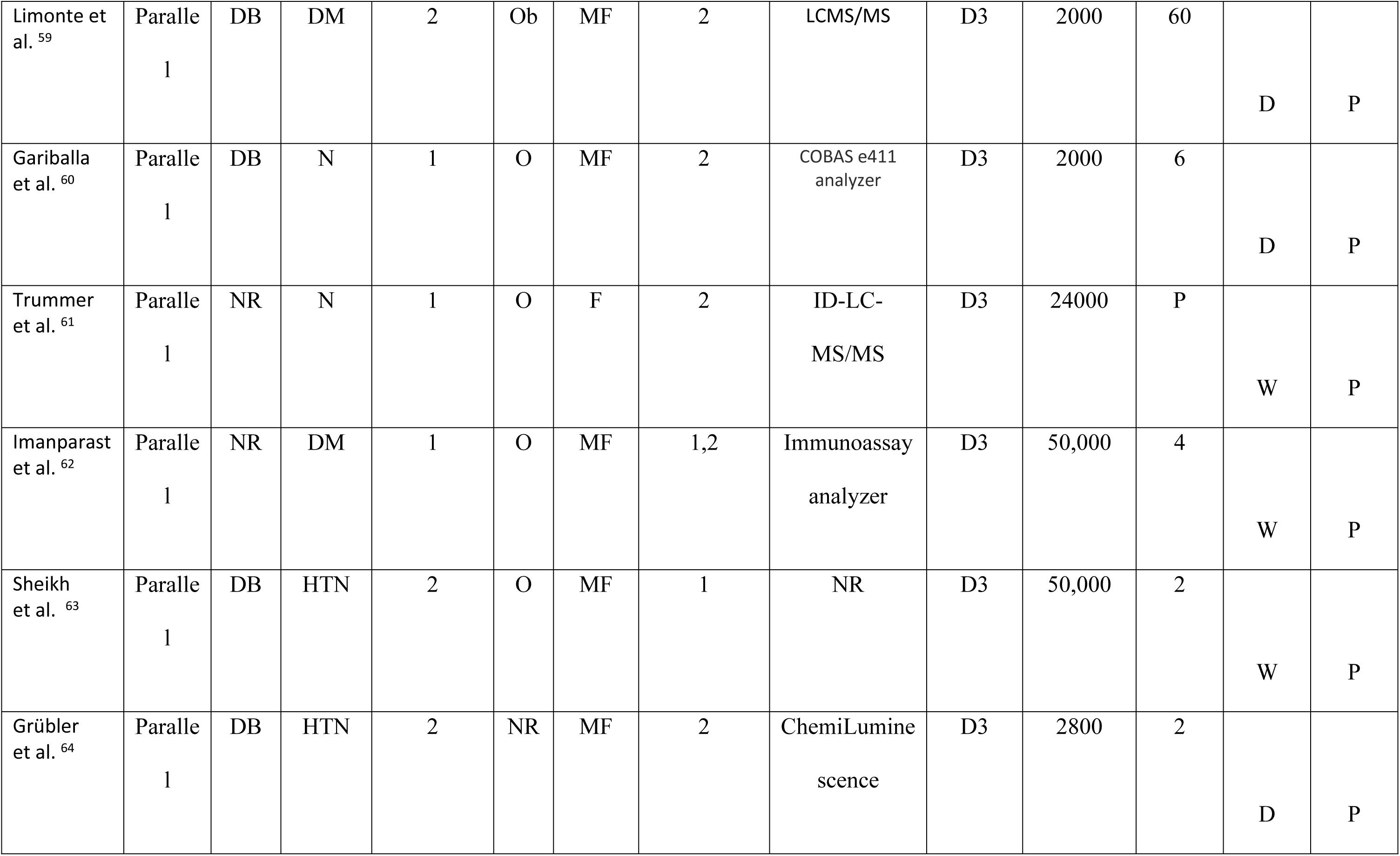
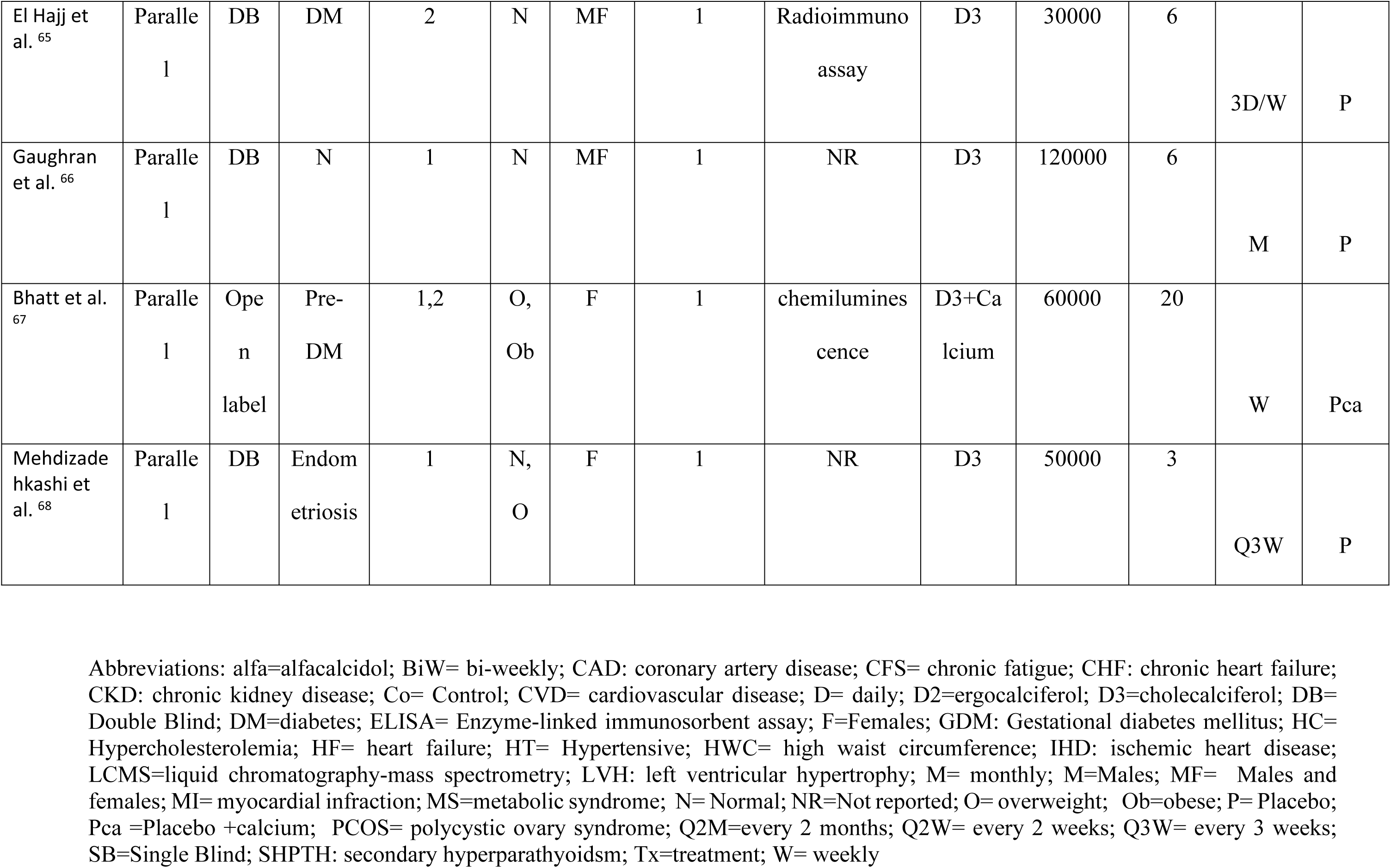
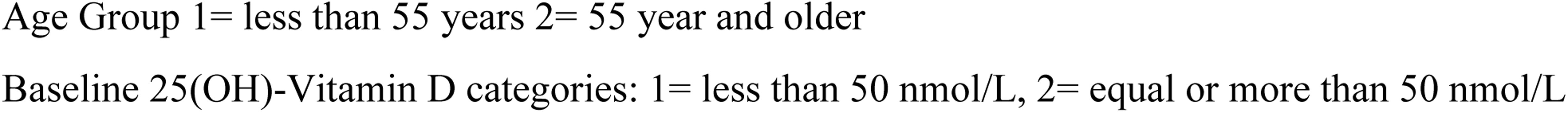
Design and participants’ characteristics of randomized, controlled studies of vitamin D supplementations and risk factors of cardiovascular disease.

As shown in Table 2, that summarizes the effect sizes (95%CI), the primary analysis reveals that compared with placebo supplementation, vitamin D did not improve lipid profile, inflammatory markers, glucose parameters, BMI. Compared to the placebo, vitamin D supplementation was associated with a modest but statistically significant reduction in systolic blood pressure (−2.80 mm Hg; 95% CI: −4.65 to −0.94), whereas no significant effect was observed for diastolic blood pressure. Considerable statistical heterogeneity was observed for most outcomes (I² >75%), indicating substantial between-study variability.

**Table 2:**
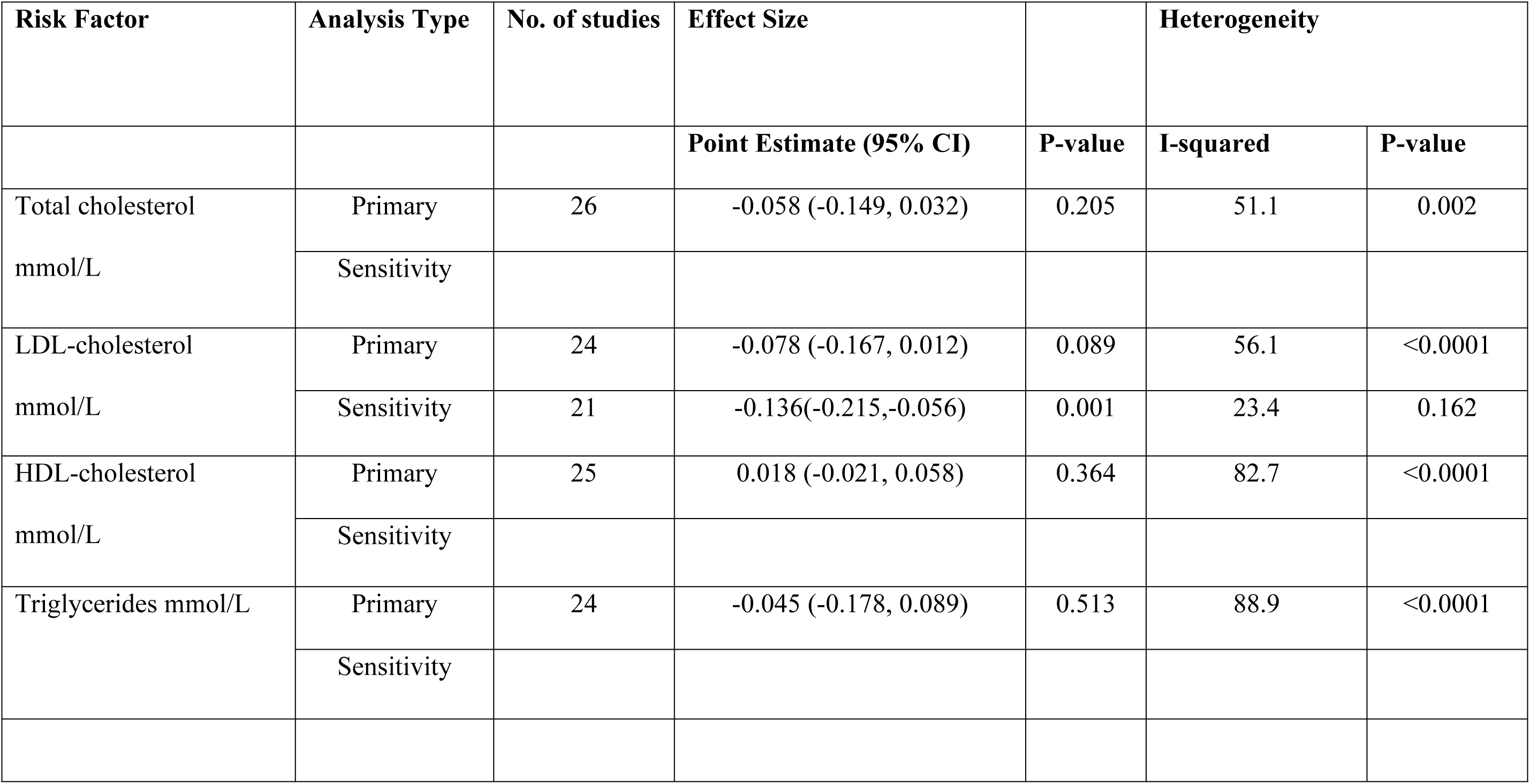
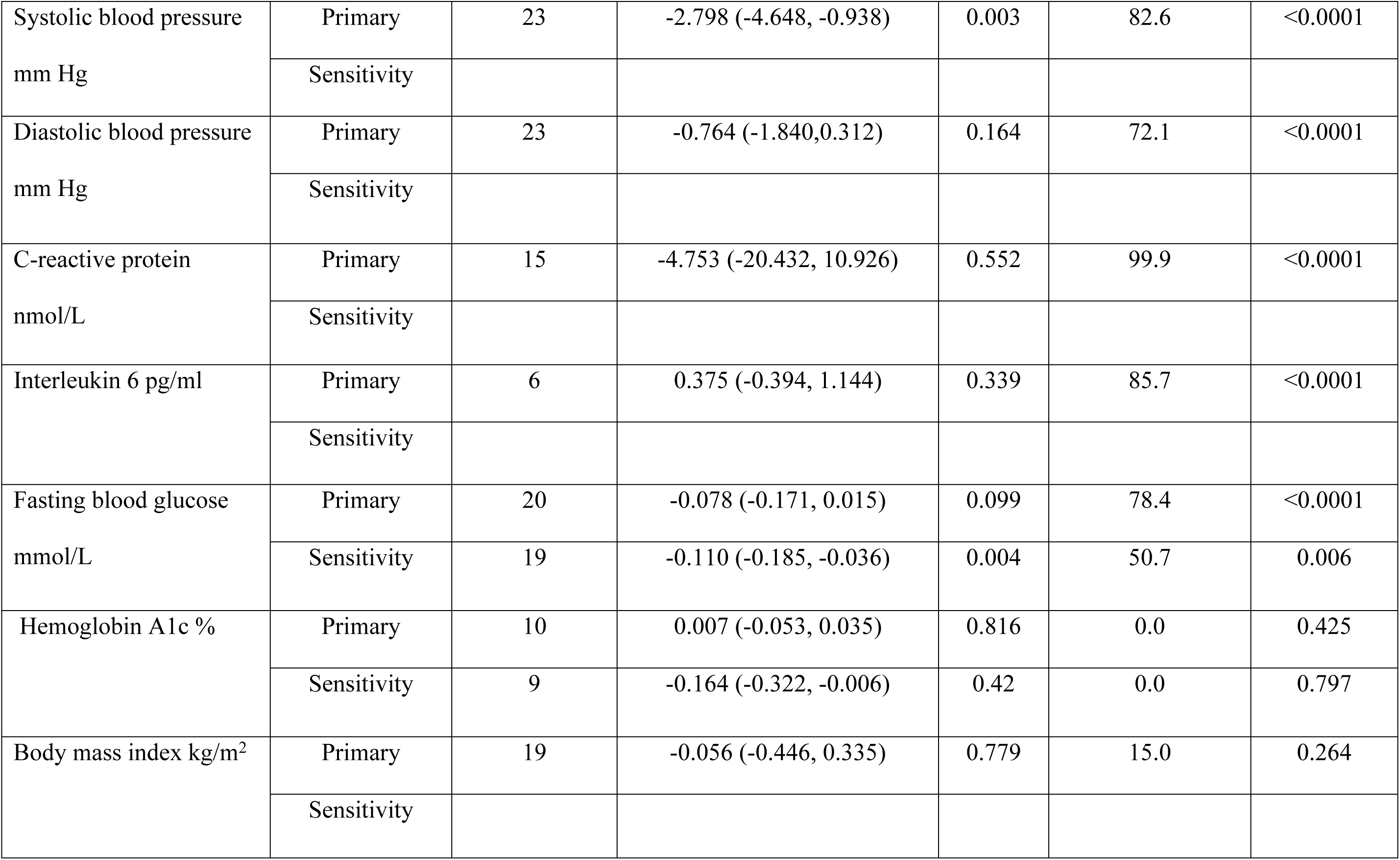
Pooled effect of clinical trials of vitamin D supplementation on cardiovascular risk factors as estimated from the primary and sensitivity analyses.

### Sensitivity Analysis

To address the considerable heterogeneity identified in the primary analysis, we conducted a series of sensitivity analyses by excluding influential trials that their effect size deviated substantially from the common distribution of the other included trials, disproportionately inflating the I^2^ values Supplementary Figures S1 and S2.

For LDL-cholesterol, exclusion of three influential trials (Kubiak *et al*; Rashad *et al*; Imanparast *et al*), all assessed as having a high-risk trial, reduced heterogeneity substantially (I² from 56.1% to 23.4%). The revised pooled estimate showed an overall significant reduction in LDL-cholesterol (−0.136 mmol/L; 95% CI: −0.215 to −0.056).

For glycemic outcomes, removal of the Kubiak *et al* trial significantly improved the findings for fasting blood glucose and hemoglobin A1c. The analysis for fasting blood glucose yielded a significant reduction (−0.110 mmol/L; 95% CI: −0.185 to −0.036) and a moderated heterogeneity (I² = 50.9%). Vitamin D supplementation reduced hemoglobin A1c by 0.164 (95%CI:-0.322, -0.006) compared with placebo with no heterogeneity observed.

For other outcomes, sensitivity analyses did not identify influential studies, and pooled estimates remained consistent with the primary analysis.

### Subgroup analysis

Subgroup analysis by age showed that reductions of LDL-cholesterol (-0.142 mmol/L, 95%CI: -0.180 to -0.105) (Figure 2a), and SBP (-3.150 mm Hg, 95%CI: -3.982 to -2.318) (Figure 2b), were significant among participants aged ≥55 years. In contrast, fasting blood glucose reduction was significant (-0.142 mmol/L, 95%CI: -0.254 to -0.03) in participants who aged less than 55 years (Figure2c).

**Figure 2.**
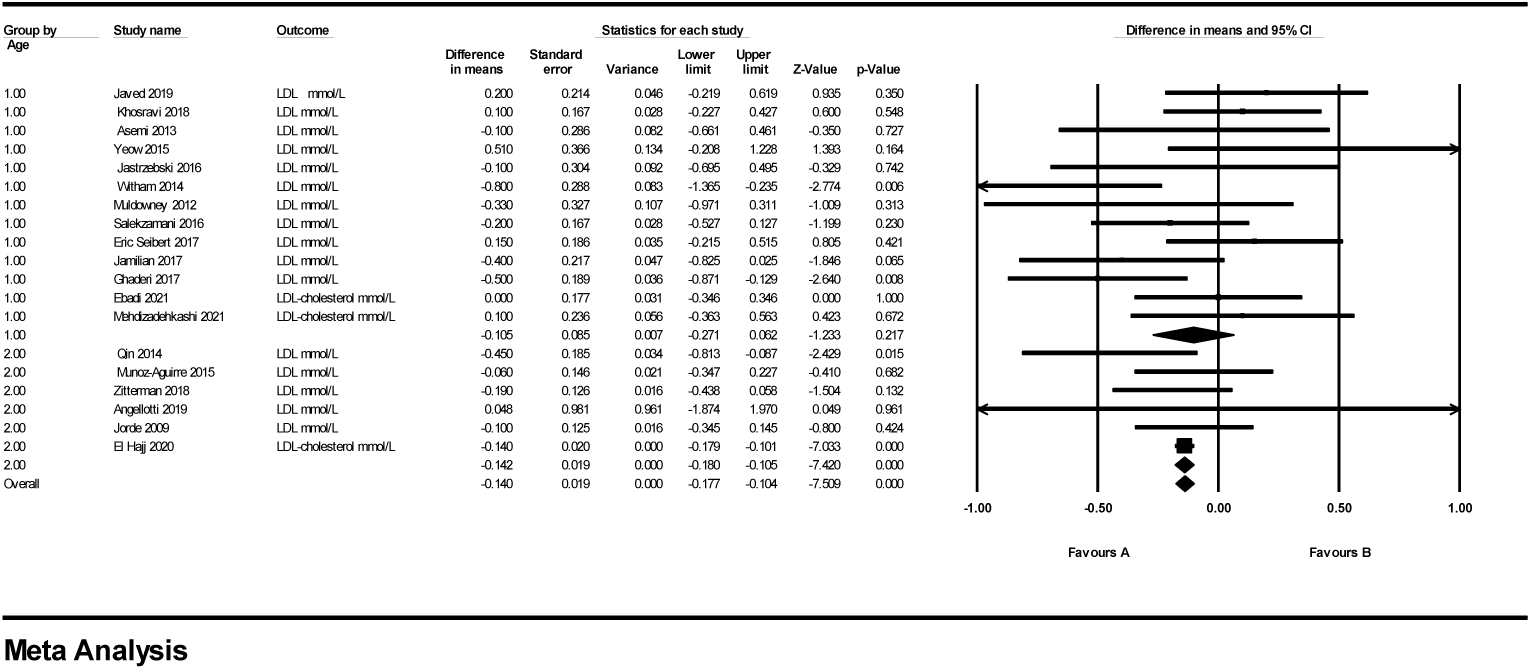
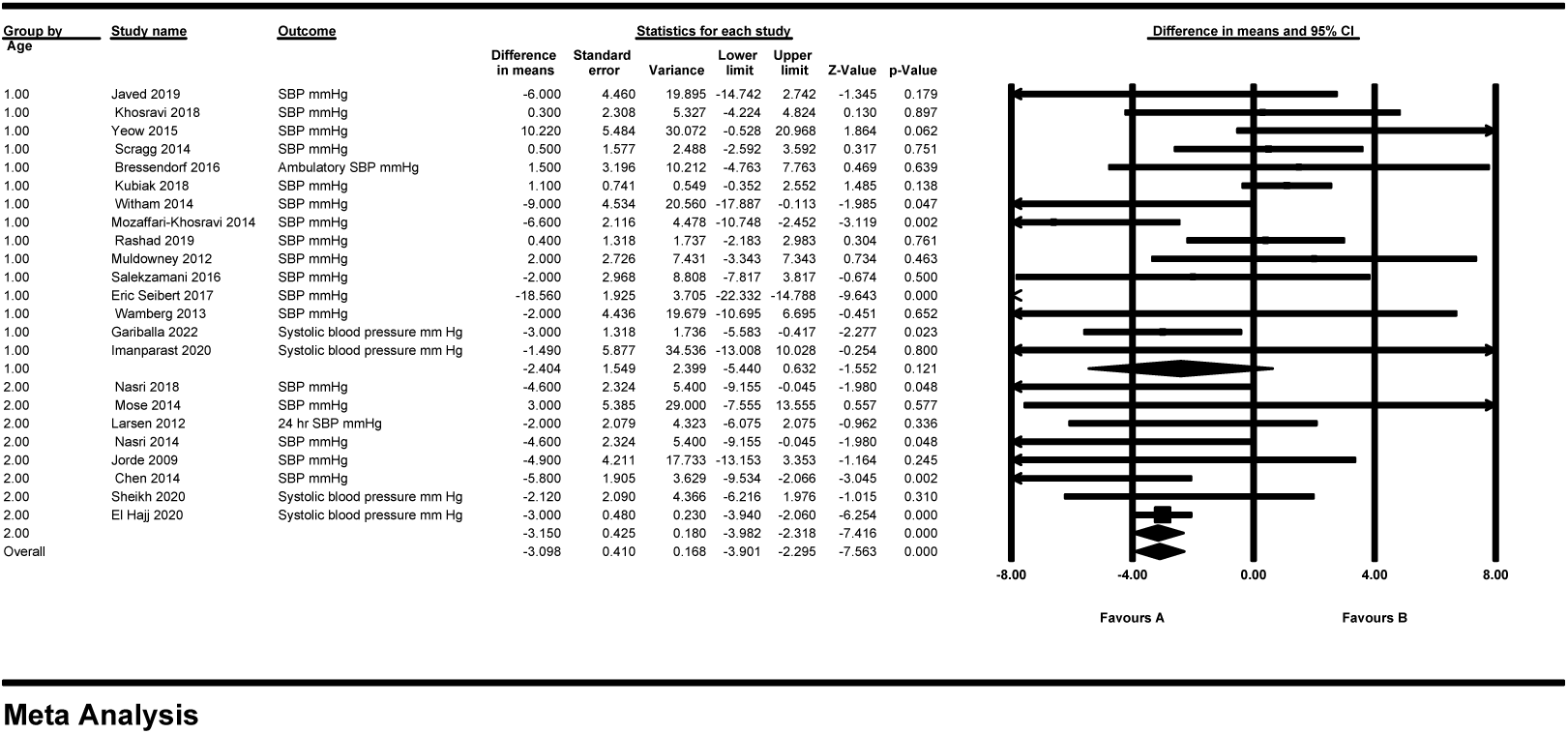
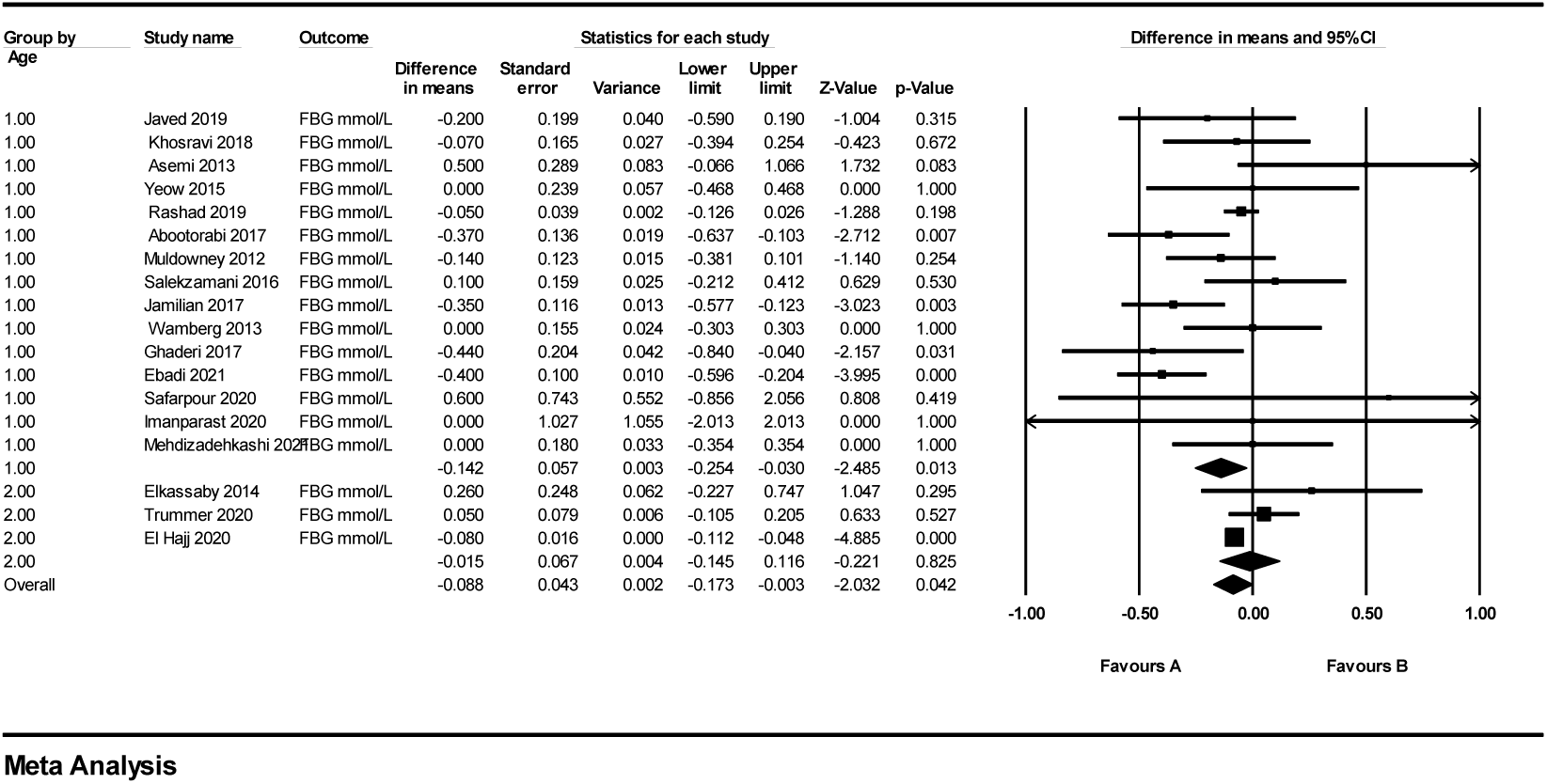
Forest plot of subgroup analysis by age of: a) low-density-lipoprotein cholesterol, b) systolic blood pressure, c) fasting blood glucose

Subgroup analyses by baseline vitamin D status showed a favorable effect on fasting blood glucose (-0.127 mmol/L, 95% CI: -0.204 to -0.050) (Figure 3a) and hemoglobin A1c (-0.252 %, 95%CI: --0.464to -0.0.039) (Figure3b).

**Figure 3.**
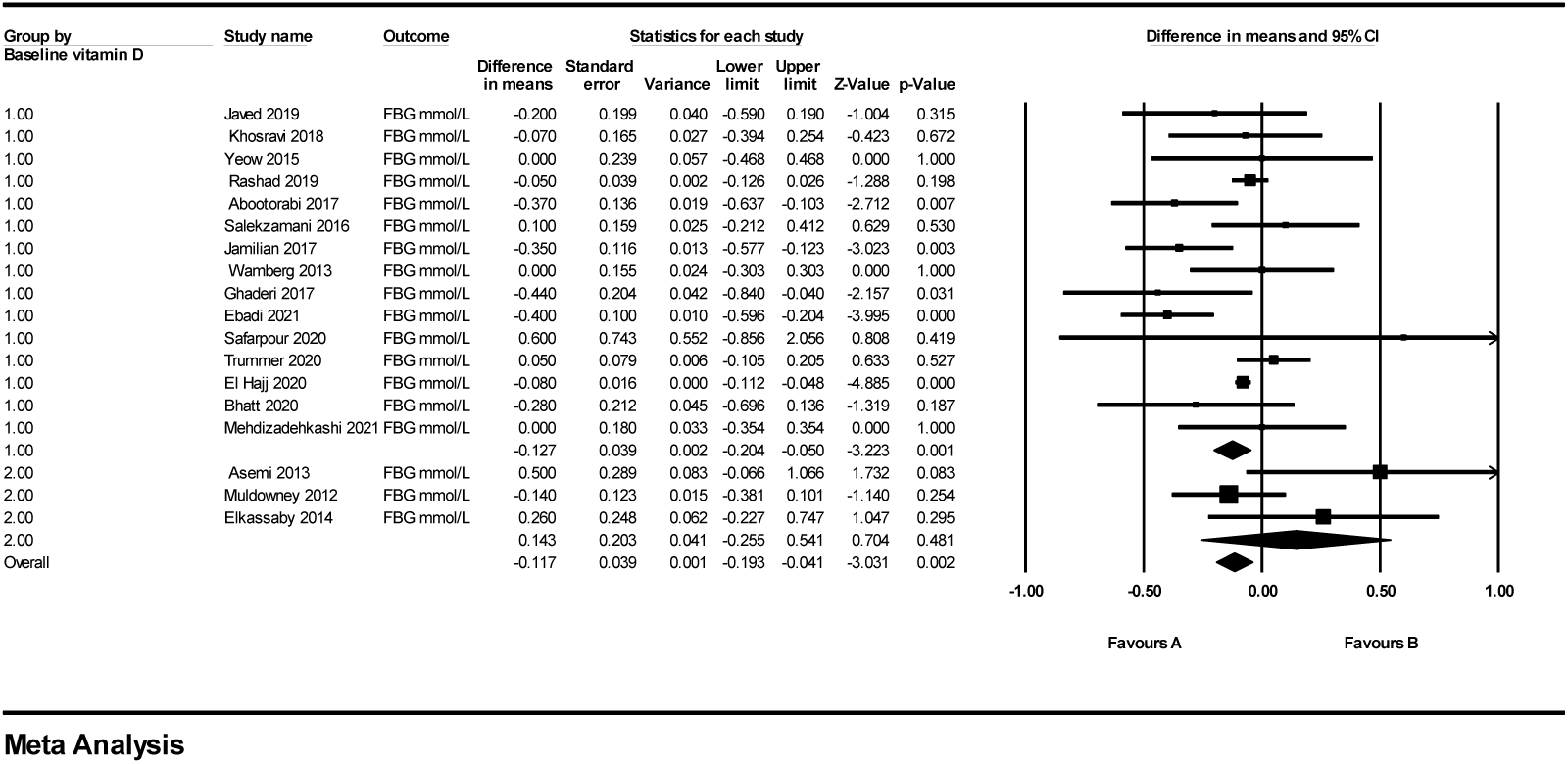
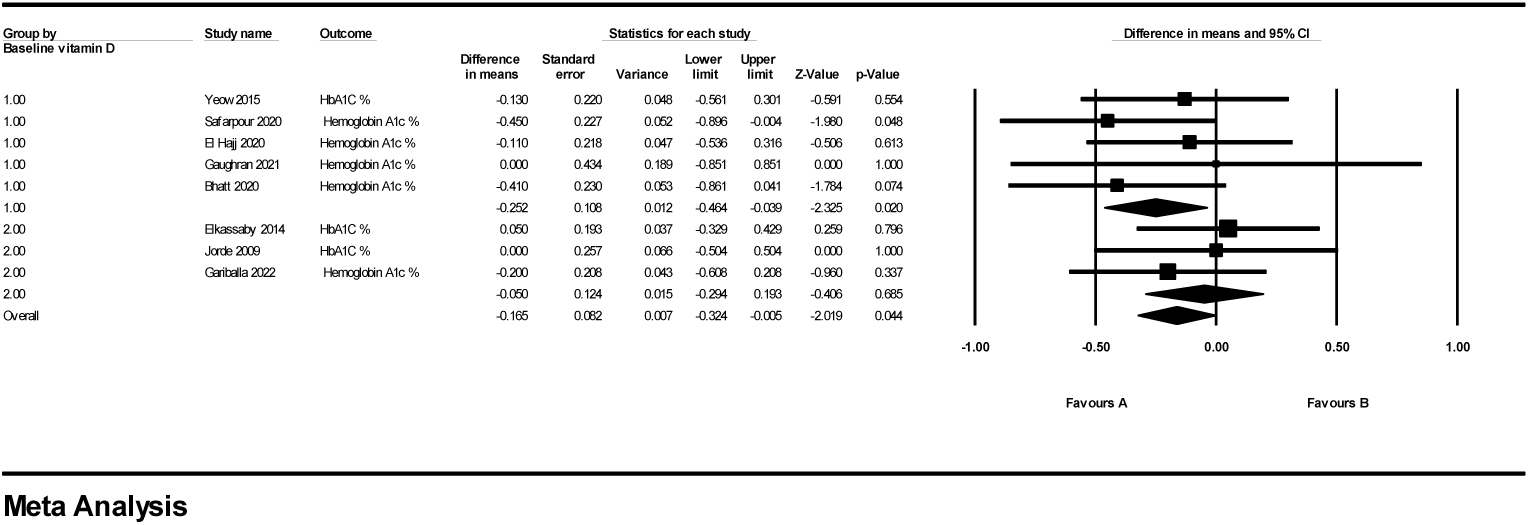
Forest plot of subgroup analysis by baseline blood vitamin D concentrations of a) fasting blood glucose, b) hemoglobin A1c

Subgroup analyses showed no significant differences in effects for the remaining outcomes across age groups or baseline vitamin D status.

### Risk of Bias and Publication Bias

A summary of each risk of bias item presented as percentages across all included studies is provided in Supplementary Figure S3. The authors’ judgment of each risk of bias item for individual studies is summarized in Supplementary Figure S4. The method of random sequence generation was performed in about 70% of the trials while allocation concealment was reported in about 30%. Around 70% of studies were judged to have an unclear risk of detection bias due to insufficient reporting of outcome assessor blinding, approximately 25% of the trials did not report whether or how blinding was achieved and 50% of the trials were at high risk of performance bias. About 25% of the trials did not provide enough information on withdrawals or loss to follow up to permit an evaluation of this. For about half of the trials, there was insufficient information to judge selective reporting.

Funnel plots for LDL-cholesterol, SBP, fasting blood pressure and hemoglobin A1c are shown in supplementary Figures S4-S8. Visual examination of the funnel plots shows a symmetrical appearance suggesting no evidence of publication bias for LDL-cholesterol, SBP, fasting blood pressure whereas asymmetry was observed for hemoglobin A1c.

## Discussion

Based on the available evidence, the present meta-analysis shows that vitamin D supplementation reduces LDL-cholesterol, SBP, fasting blood glucose and hemoglobin A1c. Moreover, this analysis provides evidence that age and baseline vitamin D blood concentration may influence the efficacy of vitamin D supplementation as a modulator for some cardiovascular disease metabolic risk factors. However, these results should be interpreted cautiously given the substantial heterogeneity and variable methodological quality of the included trials.

Previous meta-analyses have reported inconsistent effects of vitamin D supplementation on lipid parameters. A 2012 on 12 randomized clinical trials reported that vitamin D supplementation increased LDL-cholesterol concentration and did not significantly affect total cholesterol, HDL-cholesterol, or triglycerides.^11^ In the meta-analysis by Wang *et al*., ^11^ seven randomized controlled trials using cholecalciferol were included, with daily vitamin D supplementation doses ranging from 300 to 3,332 IU and intervention durations between 42 days and 3 years. The Wang *et al*., ^11^ primarily included non-Hispanic white, elderly participants and trials not specifically designed to assess lipid outcomes, thereby limiting generalizability. In contrast, a larger meta-analysis by Dibaba *et al*. in 2019, ^12^ including 41 trials, showed that vitamin D supplementation exerted beneficial effects on total cholesterol, LDL-cholesterol, and triglycerides concentrations but not HDL-cholesterol concentrations. ^12^ particularly among individuals with hypercholesterolemia and vitamin D insufficiency who are at high risk of cardiovascular disease. ^12^ Conversely, a more recent meta-analysis of seven trials of vitamin D supplementation in adults with metabolic syndrome reported no significant effects on any blood lipid parameter ^13^. These discrepancies likely reflect differences in inclusion criteria, baseline vitamin D status, participant health profiles, and supplementation regimens.

Regarding blood pressure, prior evidence has also been mixed. Witham *et al* reported DBP lowering, but no effects on SBP, due to vitamin D supplementation ^15^. whereas Elamin et al., analyzing 11 trials, found no reduction in SBP or DBP due to vitamin D supplementation^16^. Similarly, Shu *et al* reported that vitamin D supplementation did not affect blood pressure parameters in vitamin D-deficient participants from seven trials.^17^

In contrast to some previous findings, this meta-analysis using of randomised clinical trials showed an overall LDL-cholesterol and SBP reduction effect of vitamin D and this effect was particularly among participants aged 55 years and older. and that the lowering effect of vitamin D on fasting blood glucose was in participants less than 55 years of age. These findings are consistent with observations by Farapti *et al*, who found that age was a modulator for the hypotensive effect of vitamin D supplementation. ^18^. Nonetheless, additional well-designed randomized trials are required to establish the cause-effect of vitamin D supplementation on blood pressure, LDL-cholesterol, and glycemic control indicators and determine whether these reductions translate into clinically meaningful cardiovascular benefits. A plausible mechanism by which vitamin D can affect CVD risk factors is through its action on renin-angiotensin-aldosterone systems, parathyroid hormones, and insulin secretion and sensitivity. ^6^

Evidence regarding glucose metabolism remains inconclusive. While some previous meta-analyses combining previous trials failed to detect an overall favorable effect on glucose metabolism linked to vitamin D supplementation. ^19^ Tang *et al*. found that vitamin D supplementation decreased fasting blood glucose levels ^20^ consistent with our findings. In the present analysis, improvements in fasting blood glucose were mainly observed among participants younger than 55 years and those with baseline serum vitamin D concentrations below 50 nmol/L. Although a reduction in hemoglobin A1c was also observed, this finding should be interpreted cautiously due to evidence of publication bias. Proposed mechanisms linking vitamin D deficiency to insulin resistance include inflammation-mediated pathways^21^, however, this analysis showed that vitamin D does not affect inflammatory markers including c-reactive protein and interleukin-6.

Most included studies in this analysis reported a baseline 25(OH)-vitamin D of less than 50 nmol/L. suggesting widespread insufficiency among participants. It is possible that serum vitamin D levels must reach a threshold before measurable improvements improvement in cardiovascular risk factors could be detected. Previous meta-analysis suggests that a daily supplemental intake of vitamin D of 797 to 2519 IU for European adults, and 729, 2026, and 1229 IU for adults in North America, Asia, and the Middle East and Africa, respectively, are needed to achieve a 25(OH) vitamin D concentration of 75 nmol/L.^22^ Further research is needed to investigate the optimal dose of vitamin D needed to achieve a vitamin D blood concentration that is needed to modify the metabolic risk factors associated with cardiovascular disease. Importantly, whether the modest reductions reported in this analysis in SBP, LDL-cholesterol, fasting blood glucose, and hemoglobin A1c by vitamin D supplementation can be translated to a reduction in cardiovascular events in the general population remains uncertain and should be evaluated in trials specifically designed with clinical endpoints.

## Limitations

This analysis is not without limitations. First, limitations of our analysis arise from the characteristics of the included studies. Despite the use of random-effects models and rigorous sensitivity analyses and the exclusion of outliers, considerable statistical heterogeneity persisted for some outcomes. This residual heterogeneity is likely, stemming from the broad diversity in study protocols. Specifically, the included trials utilized a wide range of dosages and durations, across healthy participants and populations with distinct Pathophysiologies. Given that Vitamin D metabolism is highly dependent on baseline status and adiposity, these clinical variations naturally hindered a uniform effect size. Future studies should recruit participants based on pre-determined baseline blood vitamin D concentrations as well as obesity status based on other measurements and not solely on BMI.

Additionally, the methodological quality of the included studies was variable. A considerable proportion of the trials included were identified as having an unclear or high risk of bias, particularly regarding performance and detection bias. Consequently, the overall certainty of the evidence is limited. Finally, subgroup analyses for certain metabolic markers —although useful for hypothesis generation— were not powered to establish definitive effect modification and should be interpreted with caution until confirmed by large-scale, prospective trials specifically designed to compare the effect of age and baseline blood vitamin D concentration as potential modulators to vitamin D supplementation for risk factors associated with CVD.

## Conclusion

Based on the findings of this meta-analysis vitamin D supplementation may be associated with modest modifications in selected cardiometabolic risk factors, including systolic blood pressure, low-density lipoprotein cholesterol, fasting blood glucose, and hemoglobin A1c. Although the primary analyses exhibited substantial statistical heterogeneity, the application of robust sensitivity analyses—excluding influential outliers and studies with a high risk of bias—confirmed significant reductions in LDL-cholesterol, SBP, fasting blood glucose, and hemoglobin A1c. Consequently, the certainty and clinical relevance of these findings remain limited. Future clinical trials employing standardized protocol for vitamin D dosing, intervention durations, and conducted on participants with defined health/disease status are necessary to determine whether vitamin D supplementation confers meaningful cardiometabolic or cardiovascular benefits.

## Data Availability

All relevant data are within the manuscript and its Supporting Information files.

## Declarations

### Ethics approval and consent to participate

Ethical approval is not applicable to this article as no new data was collected.

### Consent for publication

Not applicable.

### Availability of data and materials

The data that supports the findings of this study are available on request from the corresponding author. The data are not publicly available due to privacy or ethical restrictions.

### Authors’ contributions

**Concept and design:** Suhad Abumweis, Stephanie Jew.

**Data collection:** Suhad Abumweis, Stephanie Jew, Sarah Alqadi, Ala’a Al Tarteer, Waed Alrefai, Taima Qudah.

**Analysis and interpretation:** Suhad Abumweis, Foad Alzoughool, Stephanie Jew.

**Writing the article:** Suhad Abumweis, Stephanie Jew, Waed Alrefai, Foad Alzoughool, Taima Qudah.

**Critical revision of the article:** Suhad Abumweis, Stephanie Jew.

**Final approval of the article:** All authors.

**Overall responsibility:** Suhad Abumweis, Taima Qudah.

## Abbreviations

CVD: cardiovascular disease:
LDL-cholesterol: low-density lipoprotein cholesterol
HDL-cholesterol: high-density lipoprotein cholesterol
SBP: systolic blood pressure
DBP: diastolic blood pressure
BMI: body mass index
Vitamin D3: cholecalciferol
Vitamin D2: ergocalciferol

## Acknowledgements

Not applicable.

## Supplementary Materials

- **Figure S1**. Forest plots of leave-one-out sensitivity analyses evaluating the effect of vitamin D supplementation on: (a) total cholesterol, (b) LDL-cholesterol, (c) LDL-cholesterol after exclusion of three influential studies (Kubiak *et al*.; Rashad *et al*.; Imanparast *et al*.), (d) HDL-cholesterol, (e) triglycerides, (f) systolic blood pressure, and (g) diastolic blood pressure.
- **Figure S2.** Forest plots of leave-one-out sensitivity analyses evaluating the effect of vitamin D supplementation on: (a) C-reactive protein, (b) interleukin-6, (c) fasting blood glucose, (d) fasting blood glucose after exclusion of the influential study by Kubiak et al. (2018), (e) hemoglobin A1c, (f) hemoglobin A1c after exclusion of the influential study by Kubiak et al. (2018), and (g) body mass index.
- **Figure S3:** Risk of bias graph showing review authors’ judgments about each risk of bias item, presented as percentages across all included studies.
- **Figure S4**: Risk of bias summary presenting review authors’ judgment of each risk of bias item for each included study.
- **Figure S5:** Funnel plot of standard error versus difference in means for LDL cholesterol concentrations.
- **Figure S6:** Funnel plot of standard error versus difference in means for systolic blood pressure.
- **Figure S7:** Funnel plot of standard error versus difference in means for fasting blood glucose.
- **Figure S8:** Funnel plot of standard error versus difference in means for hemoglobin A1c.

